# Pre-test probability for SARS-Cov-2-related Infection Score: the PARIS score

**DOI:** 10.1101/2020.04.28.20081687

**Authors:** On Behalf of the APHP/Universities/Inserm COVID-19 Research collaboration, Tordjman Mickael, Mekki Ahmed, Mali D. Rahul, Saab Ines, Chassagnon Guillaume, Guillo Enora, Burns Robert, Eshagh Deborah, Beaune Sebastien, Madelin Guillaume, Bessis Simon, Feydy Antoine, Mihoubi Fadila, Doumenc Benoit, Carlier Robert-Yves, Drape Jean Luc, Revel Marie-Pierre

**Affiliations:** Department of Radiology, Cochin Hospital, APHP, France; Department of Radiology, Ambroise Paré Hospital, APHP, France; Center for Biomedical Imaging, Department of Radiology, New York University School of Medicine, New York, USA; Université de Paris; Department of Internal Medicine, Saint Antoine Hospital, APHP, France; Emergency Department, Ambroise Paré Hospital, APHP, France; Department of Infectious diseases, Raymond Poincaré Hospital, APHP, France; Emergency Department, Cochin Hospital, APHP, France; Department of Radiology, Raymond Poincaré Hospital, APHP, France; DMU Smart Imaging

## Abstract

**Background:** Diagnostic tests for SARS-CoV-2 infection (mostly RT-PCR and Computed Tomography) are not widely available in numerous countries, expensive and with imperfect performance

**Methods:** This multicenter retrospective study aimed to determine a pre-test probability score for SARS-CoV-2 infection based on clinical and biological variables. Patients were recruited from emergency and infectious disease departments and were divided into a training and a validation cohort. Demographic characteristics, clinical symptoms, and results of blood tests (complete white blood cell count, serum electrolytes and CRP) were collected. The pre-test probability score was derived from univariate analyses between patients and controls, followed by multivariate binary logistic analysis to determine the independent variables associated with SARS-CoV-2 infection. Points were assigned to each variable to create the PARIS score. ROC curve analysis determined the area under the curve (AUC).

**Findings:** One hundred subjects with clinical suspicion of SARS-CoV-2 infection were included in the training cohort, and 300 other consecutive individuals were included in the validation cohort. Low lymphocyte (<1·3 G/L), eosinophil (<0·06G/L), basophil (<0·04G/L) and neutrophil counts (<5G/L) were associated with a high probability of SARS-CoV-2 infection. No clinical variable was statistically significant. The score had a good performance in the validation cohort (AUC=0.889 (CI: [0.846–0.932]; STD=0.022) with a sensitivity and Positive Predictive Value of high-probability score of 80·3% and 92·3% respectively. Furthermore, a low-probability score excluded SARS-CoV-2 infection with a Negative Predictive Value of 99.5%.

**Interpretation:** The PARIS score based on complete white blood cell count has a good performance to categorize the pre-test probability of SARS-CoV-2 infection. It could help clinicians avoid diagnostic tests in patients with a low-probability score and conversely keep on testing individuals with high-probability score but negative RT-PCR or CT. It could prove helpful in countries with a low-availability of PCR and/or CT during the current period of pandemic.

**Funding:** None

**Putting research into context:** *Evidence before this study:* In numerous countries, large population testing is impossible due to the limited availability and costs of RT-PCR kits and CT-scan. Furthermore, false-negativity of PCR or CT as well as COVID-19 pneumonia mimickers on CT may lead to inaccurate diagnoses. Pre-test probability combining clinical and biological features has proven to be a particularly useful tool, already used in clinical practice for management of patients with a suspicion of pulmonary embolism.

*Added value of this study:* This retrospective study including 400 patients with clinical suspicion of SARS-CoV-2 infection was composed of a training and a validation cohort. The pre-test probability score (PARIS score) determines 3 levels of probability of SARS-CoV2 infection based on white blood cell count (lymphocyte, eosinophil, basophil and neutrophil cell count).

*Implications of the available evidence:* This pre-test probability may help to adapt SARS-CoV-2 infection diagnostic tests. The high negative predictive value (99·5%) of the low probability category may help avoid further tests, especially during a pandemic with overwhelmed resources. A high probability score combined with typical CT features can be considered sufficient for diagnosis confirmation.

## Introduction

Reverse Transcription Polymerase Chain Reaction (RT-PCR) is currently the test of reference to diagnose patients with the emerging SARS-CoV-2 infection^1^. However, in numerous countries including France, RT-PCR kits are not available enough to allow for large population testing. A definitive diagnosis is essential to control this disease so diagnostic methods need to be improved. Computed Tomography (CT)-scan has a reported high sensitivity^2^ but radiological signs can be delayed after disease onset, with up to 56% CT negativity in the first 3 days of symptomatic infection^3^. Moreover, performing large scale CT scanning during the current pandemic is made difficult by the need to apply rigorous disinfection protocols between patients^4^. The Fleischner Society recently published a Multinational Consensus Statement on the use of thoracic imaging based on different scenarios^5^. Imaging is indicated in case of moderate to severe disease manifestations, but not indicated in case of mild symptoms consistent with COVID-19 and no risk factor for disease progression. Even though CT-scan has a higher sensitivity than PCR (71–95% for PCR and 97–98% for CT), radiologists may still experience some difficulties for differentiating COVID-19 from non-COVID pneumonia^6,7^. False-negativity of PCR or CT as well as COVID-19 pneumonia mimickers on CT may lead to inaccurate diagnoses. Pre-test probability combining clinical and biological features has proven to be a very useful tool, already used in clinical practice for management of patients with a suspicion of pulmonary embolism^8^. Therefore, we aimed to derive a new prediction score based on clinical and biological variables, independently from subjective clinical evaluation by assessing a retrospective cohort of patients admitted to the emergency department (ED) of our institution for a clinical suspicion of SARS-Cov-2 infection. We validated this **P**re-test probability for S**AR**S-Cov-2 **I**nfection based on **S**coring (PARIS score) in a distinct multicenter cohort of patients admitted for the same reason in ED or infectious disease departments of 3 hospitals.

## Methods

This retrospective observational study has been approved by our local ethic committee (Institutional Review Board N° AAA-2020–08014), which waived the need for patient consent.

### Derivation cohort

We elaborated a diagnostic strategy for SARS-Cov-2 infection based on clinical and/or biological features to determine pre-test probability before RT-PCR or CT-scan. We decided to recruit a retrospective cohort of 100 patients with both RT-PCR and CT-scan results available with a 1:1 patient:control inclusion ratio from ED at Cochin Hospital (Paris, France) with a suspicion of SARS-Cov-2 infection: 50 consecutive infected patients and 50 consecutive controls. Exclusion criteria were: 1) absence of confirmed diagnosis (diagnosis still under investigation) (N=4); 2) lack of blood test including complete white blood cell count and serum electrolytes (N=6); 3) absence of reported clinical characteristics (N=2). Between March 15^th^ and April 5^th^, 112 patients were screened for a clinically suspected SARS-Cov-2 infection. One hundred patients were finally included. Clinical examination was standardized at the ED. Demographic characteristics and comorbidities including high blood pressure, respiratory diseases (asthma, restrictive or chronic obstructive pulmonary disease (COPD)), immunodeficiency, kidney failure were recorded. Furthermore, clinical symptoms such as cough, fever, headache, diarrhea, anosmia, ageusia, oxygen desaturation were evaluated. Finally, biological tests including results of complete white blood cell count, serum electrolytes and C Reactive Protein (CRP) were analyzed. A final diagnosis of SARS-Cov-2 infection was retained if the patient had a confirmed diagnosis with both positive RT-PCR and CT-scan showing signs of COVID-19 pneumonia (usually bilateral and peripheral ground-glass and consolidated pulmonary opacities): since no test had a perfect diagnostic performance at the time of the study, we used in the derivation cohort a strict definition of patients and controls. Thus, controls were patients with both negative RT-PCR results and negative CT-scan (normal or with alternative diagnosis). We evaluated clinical and biological variables to find the ones correlated with SARS-Cov-2 infection. We only included basic biological tests, widely available and affordable, which are adapted for a pre-test probability score.

### Validation cohort

Our pre-test diagnostic score was validated on 300 consecutive subjects suspected of SARS-Cov-2 infection (different from those of the derivation cohort) with both RT-PCR and CT-scan results available, recruited from the ED and infectious disease departments of three different hospitals (Cochin Hospital, Paris; Ambroise Paré Hospital, Boulogne; Raymond Poincaré Hospital, Garches). Patients were confirmed to have SARS-Cov-2 infection when the RT-PCR result was positive. If the RT-PCR was negative, patients were considered as COVID+ in this cohort if CT-scan images evaluated by a senior radiologist specialized in thoracic imaging (>10 years of experience) were typical for COVID-19 pneumonia (N=19). The exclusion criterion was the unavailability of initial complete white blood cell count (N=9 of the 309 initially included patients).

### Statistical analysis

Univariate analyses were performed to select potential independent variables of SARS-Cov-2 infection to include in the multivariate analysis. The Mann Whitney U test was performed for comparison of continuous variables such as hemoglobin, and chi-square test was used to comparison of nominal variables. Exact p-values were computed for the Mann Whitney tests without any assumption of normality or asymptomatic approximation. A p-value of less than 0·05 was considered as statistically significant (two-tailed). The statistically significant variables were evaluated to find the optimal cut-offs to differentiate patients and controls based on receiver operating characteristic (ROC) curve analysis (upper left point of the graph) ^9^ (Supplementary Table S1). A binary logistic regression analysis was performed to measure the relationship between the categorial variable representing presence or absence of SARS-Cov-2 infection and the potential independent variables. Variables with non-statistical significance in the multivariate models were removed. Odds-Ratio (OR) were used to assign points for the pre-test probability score to the highest values (OR>10). The Hosmer-Lemeshow goodness of fit statistic^10^ and Nagelkerke’s R^2^ were used to assess the model validity^11^. We computed the pre-test probability score for each patient in the derivation cohort. We performed a second ROC curve analysis to determine the area under the curve (AUC) and to choose the cutoff values, in order to determine the low-probability group (<5% probability of SARS-Cov-2 infection), the intermediate probability-group and the high-probability group (>90% of probability of SARS-Cov-2 infection). The proportion of patients with SARS-Cov-2 infection in each group was evaluated to check the accuracy of the PARIS score.

We assessed the ability of the score to differentiate patients and controls in the validation cohort by a ROC curve analysis and computed the ROC curve. The prevalence of SARS-Cov-2 infection was calculated in the 3 probability groups (low, intermediate, and high probability score). SPSS software (version 24, Chicago, IL, USA) was used for statistical analysis.

## Results

### Derivation cohort

The clinical and biological characteristics of the patients and controls included in the derivation cohort are presented in Table 1.

**Table 1:** Characteristics of patients with SARS-CoV-2 infection (confirmed with both PCR and CT) and controls. QPulmonary disease: asthma, COPD or restrictive syndrome; Results are presented as means [standard deviation]

|  | **Patients** | **Controls** | **p-value** |
| --- | --- | --- | --- |
| n | 50 | 50 | - |
| Age | 60.8 [18.1] | 54.1 [18.6] | 0.07 |
| Sex (M :F) | 30 :20 | 25 :25 | 0.23 |
| *Clinical characteristics* |  |  |  |
| Cough | 43 (86%) | 39 (78%) | 0.30 |
| Fever | 46 (92%) | 32 (64%) | 0.001 |
| Temperature at ER | 37.8 [0.81] | 37.4 [0.96] | 0.03 |
| Shortness of breath | 35 (70%) | 31 (62%) | 0.40 |
| Saturation | 94% | 96% | 0.04 |
| Diarrhea | 12 (24%) | 6 (12%) | 0.12 |
| Myalgia | 20 (40%) | 7 (14%) | 0.003 |
| Headaches | 8 (16%) | 14 (28%) | 0.15 |
| Anosmia | 5 (10%) | 1 (2%) | 0.09 |
| Ageusia | 5 (10%) | 0 | 0.09 |
| Time from onset | 6.4d | 4.8d | 0.09 |
| *Comorbidities* |  |  |  |
| Comorbidities | 29 (58%) | 35 (70%) | 0.21 |
| Diabetes | 5 (10%) | 8 (16%) | 0.38 |
| Hypertension | 14 (28%) | 10 (20%) | 0.35 |
| Renal failure | 2 (4%) | 2 (4%) | 1 |
| Pulmonary disease<sup>⌘</sup> | 9 (18%) | 18 (36%) | 0.04 |
| Immunodeficiency | 7 (14%) | 16 (32%) | 0.03 |
| *Blood tests* |  |  |  |
| Hemoglobin | 13.6 [1.9] | 13.4 [1.6] | 0.61 |
| Lymphocytes | 0.93 [0.47] | 2.02 [0.92] | <10<sup>-3</sup> |
| Neutrophils | 4.58 [2.69] | 7.24 [4.50] | <10<sup>-3</sup> |
| Eosinophils | 0.03 [0.10] | 0.19 [0.26] | <10<sup>-3</sup> |
| Basophils | 0.02 [0.01] | 0.05 [0.04] | <10<sup>-3</sup> |
| Monocytes | 0.51 [0.33] | 0.83 [0.46] | <10<sup>-3</sup> |
| Platelets | 207 [91] | 270 [102] | 0.002 |
| Sodium | 135.7 [2.7] | 136.1 [3.6] | 0.43 |
| Potassium | 4.0 [0.46] | 4.1 [0.43] | 0.46 |
| Chloride | 97.1 [3.2] | 98.0 [4.0] | 0.14 |
| Bicarbonate | 24.0 [3.2] | 23.6 [3.1] | 0.53 |
| Total protein | 71.8 [5.5] | 74.0 [6.3] | 0.09 |
| Creatinine | 95.6 [65.8] | 84.0 [28.1] | 0.54 |
| CRP | 58.8 [45.9] | 64.6 [116.4] | 0.29 |

Fever/temperature at ER, oxygen saturation and myalgias were the clinical variables that significantly differed between patients and controls. Patients presented with significantly lower frequency of pulmonary diseases (asthma, COPD or restrictive lung disease) and immunodeficiency. Among blood test variables, we noted significantly lower lymphocyte, neutrophil, eosinophil, monocyte, and basophil cell counts in patients compared to controls. Additionally, platelet count was significantly lower in patients with SARS-Cov-2 infection. No difference between the 2 groups was found for serum electrolytes and CRP.

All these variables were included in the multivariate binary logistic regression analysis. Only low lymphocyte, basophil, eosinophil, and neutrophil cell counts were significantly associated with SARS-Cov-2 infection (Table 2). ROC curve analysis determined the best cut-offs for sensitivity and specificity (Supplementary Table 1). Points were assigned according to the odds-ratio: only lymphopenia (<1·3G/L) was associated with OR>10 (12·7) and 2 points were consequently assigned. The final score is presented in Table 3. The score was evaluated in the cohort of derivation (AUC=0·921; STD=0·027; CI= [0·867–0·974]).

**Table 2:** Binary logistic regression using descending Wald model; OR: Odd-Ratio; Nagelkerke R^2^=0.74; Hosmer-Lemeshow goodness of fit statistic: p=0.63

|  | Patients | Controls | B | Exp(B)/OR | p-value |
| --- | --- | --- | --- | --- | --- |
| Basophils<4G/L | 46 (92%) | 14 (28%) | 1.66 | 5.27 | 0.03 |
| Eosinophils <0.06G/L | 44 (88%) | 16 (32%) | 1.98 | 7.23 | 0.01 |
| Lymphocytes<1.3G/L | 44 (88%) | 10 (20%) | 2.54 | 12.72 | 0.001 |
| Neutrophils<5G/L | 37 (74%) | 17 (34%) | 2.10 | 8.17 | 0.006 |

**Table 3:** Pre-test diagnostic probability of COVID-19 infection: PARIS score

| Variables | Points |
| --- | --- |
| Eosinophils < 0.06 G/L | 1 |
| Lymphocytes < 1.3 G/L | 2 |
| Neutrophils <5G/L | 1 |
| Basophils <0.04G/L | 1 |
| Score = 0-1 → Low probability |  |
| Score= 2-3 → Intermediate probability |  |
| Score ≥4 → High probability |  |

### Validation cohort

Individuals included in the validation cohort had a mean age of 63 years (significantly older than those of the training cohort; p=0·007), with 172 being males and 128 females. There were 208 SARS-Cov-2 infected patients (69·3%) and 92 controls (30·7%). The pre-test probability score had a good performance in this cohort (AUC=0·889 (CI: [0·846–0·932]; STD=0·022). The sensitivity and specificity of the high-probability score (score 4 or 5) were 80.3% and 84.8% respectively (Table 4). Furthermore, Negative Predictive Value (NPV) of a low-probability score (score 0 or 1) was 99·5%. A low-probability score included 48·9% of controls (N=45/92). Many controls (N=33; 35·9%) presented with intermediate probability scores (score 2 to 4) but the NPV of combined low- and intermediate-probability scores was only 65·5%. Forty of the 208 SARS-Cov-2 infected patients (19·2%) from the cohort of validation had an intermediate score. No patient with SARS-Cov-2 infection had a score equal to 0 and there was only one infected patient with a score of 1. Only three of the 79 individuals with a score of 5 were not infected with SARS-Cov-2 (Positive Predictive Value of 96·2%). ROC curve for the validation cohort is presented in Figure 1.

**Figure 1:**
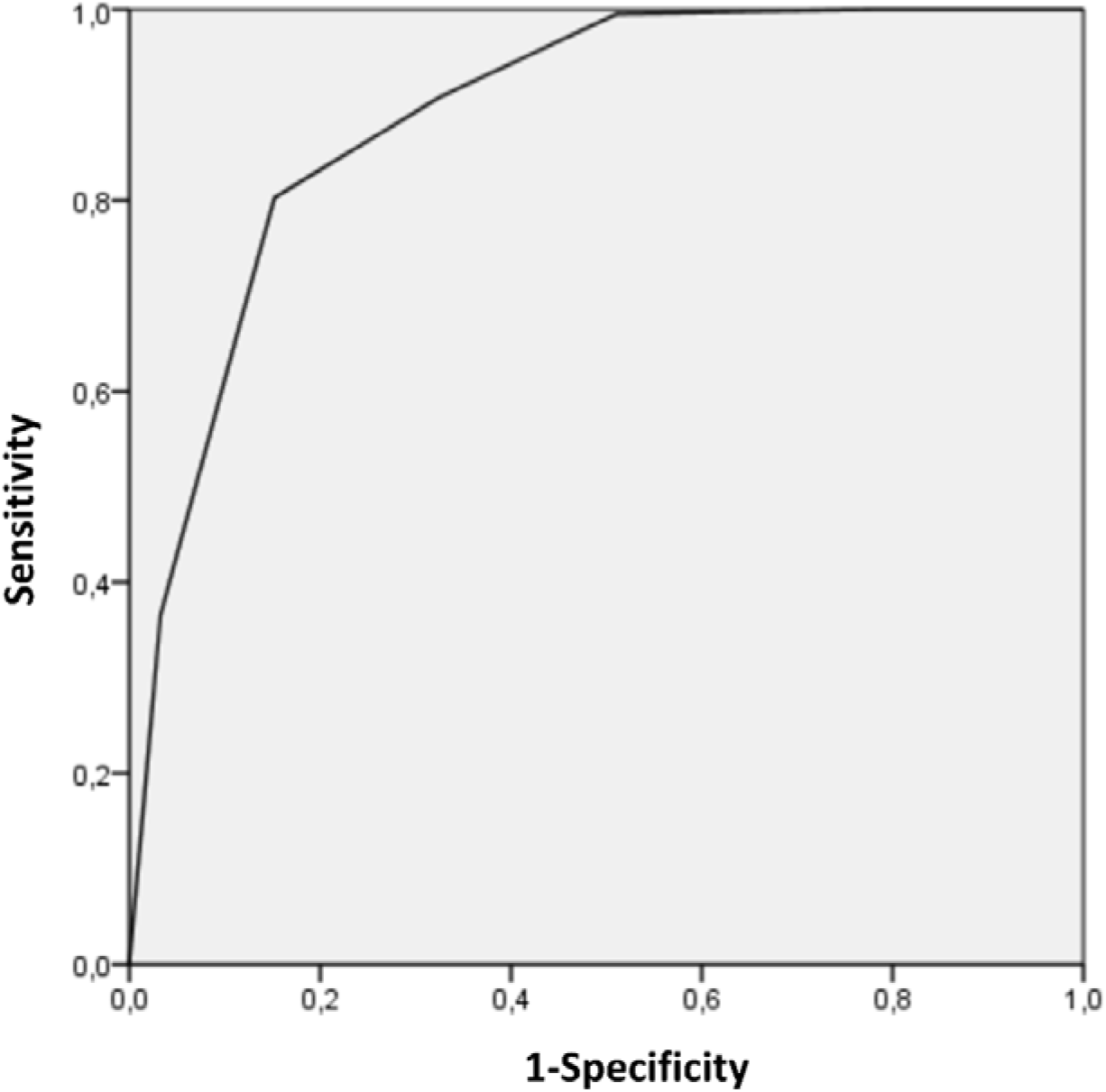
Receiver Operating Characteristic (ROC) curve of the PARIS score for the validation cohort. Area Under the Curve= 0.889

**Table 4:**
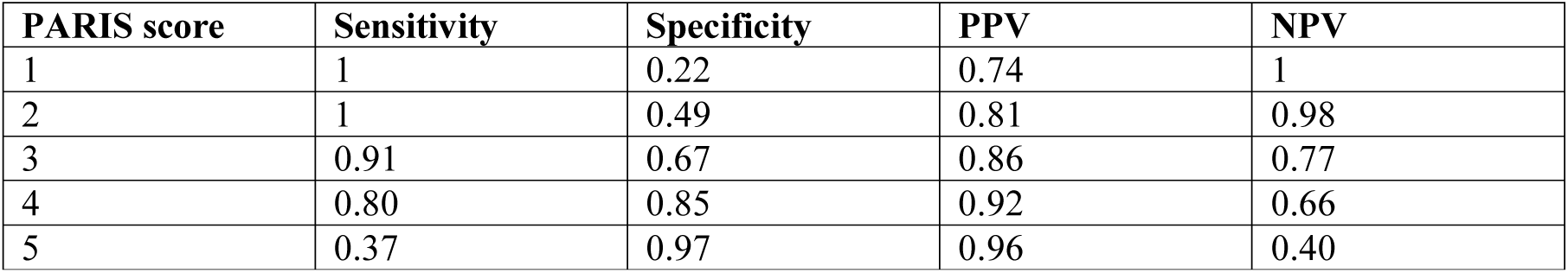
Performance in the validation cohort depending on the value of the PARIS score; PPV=Positive Predictive Value; NPV= Negative Predictive Value

| PARIS score | Sensitivity | Specificity | PPV | NPV |
| --- | --- | --- | --- | --- |
| 1 | 1 | 0.22 | 0.74 | 1 |
| 2 | 1 | 0.49 | 0.81 | 0.98 |
| 3 | 0.91 | 0.67 | 0.86 | 0.77 |
| 4 | 0.80 | 0.85 | 0.92 | 0.66 |
| 5 | 0.37 | 0.97 | 0.96 | 0.40 |

### Misclassification and reclassification

Fourteen controls presented with a high-probability score (≥4). The explanation for their leukopenia was septic shock (N=3), immunodepression induced by lupus (N=2), cystic fibrosis with lung transplantation (N=2), pancytopenia due to hematological disorder (N=2), chemotherapy for lymphoma (N=1). Four controls didn’t have any final diagnosis explaining their clinical symptoms and leukopenia and could therefore be false negative for SARS-Cov-2 infection. Only one patient was considered as a false-negative of the pre-test probability score with a low-probability score of 1. This patient had negative RT-PCR and minimal involvement on CT-scan (small ground glass opacities).

However, the score was also helpful to reclassify patients: Six out of 10 infected patients, based on RT-PCR, with negative or indeterminate CT-scan results had a high-probability score (while the 4 other patients had an intermediate PARIS score). Fifteen out of 21 patients with negative PCR but CT features typical for COVID-19 pneumonia had a high-probability score (Five other patients had an intermediate score and one patient had a low score).

## Discussion

In this study, we established a pre-test probability score (PARIS score) based on simple, affordable, and widely available blood tests (complete white blood cell count). No clinical variable was independently associated with SARS-Cov-2 infection but four biological variables(lymphocytes<1·3G/L,eosinophils<0·06G/L,basophiles<0·04G/L, neutrophils<5G/L) were associated to the condition. This finding could be particularly useful for countries with a low availability of diagnostic tests.

A low lymphocyte count has been described in patients with SARS-Cov-2 infection, possibly due to a decreased level of T-lymphocytes^12^. Furthermore, the total number of NK and CD8+ T cells has been reported to be markedly decreased in patients with SARS-Cov-2 infection^13^. Decreased absolute number of T lymphocytes is more pronounced in severe cases while decreased CD8+ T cells count could be an independent variable associated with the severity of the disease^14–16^. Eosinophils have also been reported to be decreased in previous studies^17,18^. Increasing eosinophil count may even be a predictor of improvement in patients with SARS-Cov-2 infection^19^. A low proportion of basophils has also been reported in severe cases^12^ but no previous study has reported low basophil cell count as a marker of SARS-Cov-2 infection. Furthermore, previous reports demonstrated low levels of neutrophils in SARS-Cov-2 patients^20^ but increased neutrophil count in severe cases of COVID-19 pneumonia^21^. Two previous histological studies of autopsies performed in COVID-19 patients showed inflammatory cells including lymphocytes and macrophages in the alveoli, with minimal eosinophilic and neutrophilic infiltration^22,23^. Decreased lymphocyte blood cell count could therefore be due to lymphocytic lung infiltration.

The Fleischner Society recently released a report highlighting that no score was available to assess pre-test probability in patients with mild symptoms consistent with SARS-Cov-2 infection^5^. This probability is only based on background prevalence of the disease and individual’s exposure risk. Committee members disapproved imaging use in case of mild symptoms and a negative COVID-19 test. The PARIS score may enhance the proposed Fleischner diagnostic algorithm by improving the pre-test performance. Thus, in case of high pre-test probability with no comorbidities explaining leukopenia, a second RT-PCR test or an additional CT-scan may be useful to establish the final diagnosis. Furthermore, a low PARIS score may obviate the need for RT-PCR or CT-scan, especially in a period of diagnostic test shortage.

A previous review indicated that prediction models for diagnosis and prognosis of COVID-19 infection are poorly reported, at high risk of bias, and with reported performance probably too optimistic^24^. We report here a multicenter study assessing diagnostic pre-test probability with a good accuracy, using a study design similar to that of the revised Geneve Score for pulmonary embolism^8^.

Clinical symptoms were not included in our model, since most of patients and controls presented with the same symptomatology, with no specificity of symptoms. Even oxygen saturation was not a discriminative feature. It could be explained by the fact that most patients with SARS-Cov2 infection were not severe, with subnormal oxygen saturation. Furthermore, even if patients with lung diseases were more frequent in the control group, their average oxygen saturation was 96%. Thus, low oxygen saturation may be an additional argument of SARS-Cov-2 infection in patients with no previous pulmonary disease. However, the PARIS score is mostly useful in patients with relatively mild clinical presentation. It could also be useful for initial triage of patients presenting to the ED because it is fast, affordable and allows patients with a low probability score to stay apart from those most likely to be infected, avoiding the risk of contamination.

Our study presents inherent limitations. First, its retrospective design may introduce a selection bias. Only 100 patients were included in the derivation cohort in order to recruit a similar number of infected patients and controls (50 in each group). There was no statistically significant difference between demographic characteristics of patients and controls but the 2 groups were not matched and controls more frequently presented pulmonary diseases and immunodeficiencies. A small number of patients was included due to the urgent need of diagnostic tools, but this is at risk of low reproducibility in a larger population. Indeed, the performance of the pre-test score was slightly lower in the validation cohort. We used only regular blood tests with complete white blood cell count, serum electrolytes and CRP, and did not evaluate LDH or D-dimers, since they seem to be mainly correlated with severity and have been described as risk factors for acute respiratory distress syndrome^25^. Another limit of this study is the only inclusion of patients from hospital wards, which probably induces a recruitment bias, since they may not be representative of patients with milder symptoms who consult their general practitioner, even though most symptomatic patients are directly referred to ED in our country. The evaluation of the performance of our pre-test probability score in a larger population would be of interest. Even though a part of patients in our cohort were admitted to Intensive Care Units (ICUs) shortly after presentation, more severe patients directly admitted in ICUs, may be underrepresented. However, the score might not be as useful for severe patients admitted to ICU, since they require admission and urgent care, regardless of RT-PCR positivity. This score was also built during a pandemic with high prevalence of the disease and might not be as efficient in a situation where SARS-Cov2 would be less circulating. Numerous diseases or conditions may mimic SARS-Cov-2 biological changes with leukopenia and lymphopenia. Clinicians should be aware of the limits of this score in case of hematological disease history. Lymphocyte count may be high in infected patients with leukemia while myelodysplasia may falsely induce a high-probability score. This was observed in a patient with typhoid fever, inducing a high probability score in the cohort of derivation.

In conclusion, the PARIS score shows good performance with high positive predictive value for intermediate or high score and high negative predictive value to exclude SARS-Cov-2 infection in low score individuals. It may help avoid useless repeated tests in case of low probability, especially during a pandemic with no possibility for large scale testing. Finally, a high score with typical CT features can be considered as sufficient to confirm the diagnosis.

## Data Availability

All data from this manuscript are available

## Acknowledgments

We acknowledge APHP, universities, Inserm for COVID-19 research collaboration. We acknowledge Dr Martin, Dr Alperin, Pr Lee, Pr Allain and all doctors from the Emergency Department, ICU and hospitalization departments at Cochin Hospital.

---

**Supplementary Table 1:** Receiver Operator characteristic (ROC) curve analyses to determine the adequate cut-off for the independent variables; AUC= Area Under the Curve

| Value (G/L) | Sensitivity | Specificity | AUC |
| --- | --- | --- | --- |
| Lymphocytes |  |  |  |
| 0.3 | 0.06 | 1 | 0.87 |
| 0.5 | 0.15 | 0.96 |  |
| 0.7 | 0.33 | 0.96 |  |
| 0.9 | 0.53 | 0.90 |  |
| 1.1 | 0.78 | 0.86 |  |
| 1.3 | 0.88 | 0.80 |  |
| 1.5 | 0.90 | 0.70 |  |
| 1.7 | 0.92 | 0.62 |  |
| 1.9 | 0.94 | 0.56 |  |
| 2.1 | 0.98 | 0.46 |  |
| Eosinophils |  |  |  |
| 0.01 | 0.71 | 0.83 | 0.82 |
| 0.02 | 0.79 | 0.81 |  |
| 0.03 | 0.84 | 0.76 |  |
| 0.04 | 0.84 | 0.70 |  |
| 0.05 | 0.86 | 0.68 |  |
| 0.06 | 0.88 | 0.66 |  |
| 0.07 | 0.90 | 0.62 |  |
| 0.08 | 0.92 | 0.59 |  |
| 0.09 | 0.92 | 0.58 |  |
| 0.10 | 0.92 | 0.53 |  |
| Basophils |  |  |  |
| 0.01 | 0.35 | 0.92 | 0.85 |
| 0.02 | 0.66 | 0.84 |  |
| 0.03 | 0.84 | 0.75 |  |
| 0.04 | 0.94 | 0.62 |  |
| 0.05 | 0.98 | 0.42 |  |
| 0.06 | 1 | 0.26 |  |
| Neutrophils |  |  |  |
| 2 | 0.12 | 0.95 | 0.72 |
| 3 | 0.38 | 0.88 |  |
| 4 | 0.54 | 0.78 |  |
| 5 | 0.74 | 0.68 |  |
| 6 | 0.76 | 0.58 |  |
| 7 | 0.82 | 0.47 |  |
| 8 | 0.84 | 0.32 |  |

## Authors’ contribution

<u>Study conception and design:</u> Tordjman, Revel

<u>Acquisition of data:</u> Mekki, Saab, Chassagnon, Guillo, Doumenc, Mihoubi, Feydy, Bessis, Beaune

<u>Analysis and interpretation of data:</u> Mali, Madelin, Tordjman, Mekki, Eshagh

<u>Drafting of manuscript:</u> Tordjman, Mekki, Saab, Mihoubi, Burns, Mali, Eshagh, Revel

<u>Critical revision:</u> Chassagnon, Mihoubi, Madelin, Eshagh, Beaune, Bessis, Doumenc, Feydy, Carlier, Drapé, Revel

All authors discussed the results and commented on the manuscript. All authors gave final approval of the version to be submitted.

